# Antibodies to SARS-CoV-2 are associated with protection against reinfection

**DOI:** 10.1101/2020.11.18.20234369

**Authors:** Sheila F Lumley, Denise O’Donnell, Nicole E Stoesser, Philippa C Matthews, Alison Howarth, Stephanie B Hatch, Brian D Marsden, Stuart Cox, Tim James, Fiona Warren, Liam J Peck, Thomas G Ritter, Zoe de Toledo, Laura Warren, David Axten, Richard J Cornall, E Yvonne Jones, David I Stuart, Gavin Screaton, Daniel Ebner, Sarah Hoosdally, Meera Chand, Oxford University Hospitals Staff Testing Group, Derrick W Crook, Anne-Marie O’Donnell, Christopher P Conlon, Koen B Pouwels, A Sarah Walker, Tim EA Peto, Susan Hopkins, Timothy M Walker, Katie Jeffery, David W Eyre

## Abstract

**Background:** It is critical to understand whether infection with Severe Acute Respiratory Syndrome Coronavirus 2 (SARS-CoV-2) protects from subsequent reinfection.

**Methods:** We investigated the incidence of SARS-CoV-2 PCR-positive results in seropositive and seronegative healthcare workers (HCWs) attending asymptomatic and symptomatic staff testing at Oxford University Hospitals, UK. Baseline antibody status was determined using anti-spike and/or anti-nucleocapsid IgG assays and staff followed for up to 30 weeks. We used Poisson regression to estimate the relative incidence of PCR-positive results and new symptomatic infection by antibody status, accounting for age, gender and changes in incidence over time.

**Results:** A total of 12219 HCWs participated and had anti-spike IgG measured, 11052 were followed up after negative and 1246 after positive antibody results including 79 who seroconverted during follow up. 89 PCR-confirmed symptomatic infections occurred in seronegative individuals (0.46 cases per 10,000 days at risk) and no symptomatic infections in those with anti-spike antibodies. Additionally, 76 (0.40/10,000 days at risk) anti-spike IgG seronegative individuals had PCR-positive tests in asymptomatic screening, compared to 3 (0.21/10,000 days at risk) seropositive individuals. Overall, positive baseline anti-spike antibodies were associated with lower rates of PCR-positivity (with or without symptoms) (adjusted rate ratio 0.24 [95%CI 0.08-0.76, p=0.015]). Rate ratios were similar using anti-nucleocapsid IgG alone or combined with anti-spike IgG to determine baseline status.

**Conclusions:** Prior SARS-CoV-2 infection that generated antibody responses offered protection from reinfection for most people in the six months following infection. Further work is required to determine the long-term duration and correlates of post-infection immunity.

## Introduction

SARS-CoV-2 infection produces detectable immune responses in most cases reported to date; however, it is uncertain whether previously infected people are protected from a second infection. Understanding whether post-infection immunity exists, how long it lasts, and the extent to which it may prevent, or reduce the severity of symptomatic reinfection, has major implications for the future of the SARS-CoV-2 pandemic and the corresponding global public health response.

Post-infection immunity may be conferred by various humoral and cell-mediated immune responses. Key considerations when investigating post-infection immunity include identifying functional correlates of protection, identifying surrogate tests (usually antibody assays) as markers that may not be protective in themselves but act as a proxy for protection, and defining the end-points of interest e.g. prevention of reinfection, disease, hospitalisation, death or onward-transmission.^1^

The assay-dependent antibody dynamics of SARS-CoV-2 anti-spike and anti-nucleocapsid antibodies are beginning to be defined.^2–6^ Neutralising antibodies against the receptor-binding domain of the spike protein may provide some post-infection immunity; however, this has yet to be demonstrated in longitudinal studies. Additionally, the association between antibody titres and plasma neutralising activity is assay-dependent and becomes weaker over time.^7–10^

Evidence for post-infection immunity is emerging. Despite an estimated 55 million people infected worldwide and high rates of ongoing transmission, reports of SARS-CoV-2 reinfection are few, mostly in individuals with mild or asymptomatic primary infection.^11–20^ Although a lack of widely-available PCR testing early in the pandemic may limit the numbers of confirmed reinfections reported, this suggests that infection with SARS-CoV-2 provides some protective immunity against reinfection in most people.

However no prospective longitudinal studies have yet compared infection rates in seropositive and seronegative individuals, however there are some small-scale reports that suggest antibodies may be associated with protection against infection.^21^

Here we present follow-up from a prospective longitudinal cohort study of healthcare workers (HCWs) at Oxford University Hospitals (OUH). Comparing the incidence of PCR-positive results and symptomatic infection in seropositive versus seronegative HCWs during up to 30 weeks of follow-up demonstrates post-infection immunity lasting at least 6 months.

## Methods

### Cohort description

OUH offers both symptomatic and asymptomatic SARS-CoV-2 staff testing programs, defining a HCW as anyone working at its four teaching hospital sites in Oxfordshire, UK.

SARS-CoV-2 PCR testing of nasal and oropharyngeal swabs for all symptomatic (new persistent cough, fever ≥37.8°C, anosmia/ageusia) staff was offered from 27-March-2020 onwards. PCR-positive results from community-based symptomatic testing of OUH HCWs forwarded by public health agencies were also included.

Asymptomatic HCWs were invited to participate in voluntary SARS-CoV-2 testing by nasal and oropharyngeal swab PCR and serological testing from 23-April-2020 onwards. The cohort, associated methods, baseline and 6-month antibody trajectory findings have been previously described.^5,22–24^ Following initial PCR and antibody testing, asymptomatic HCWs were invited to optionally attend for serological testing once every two months, with some offered more frequent screening as part of related studies. Asymptomatic staff were also offered optional SARS-CoV-2 PCR tests every two weeks.

### Laboratory assays

Serological investigations were performed using an enzyme-linked immunosorbent assay platform developed by the University of Oxford detecting IgG to SARS-CoV-2 trimeric spike antigen, using net-normalised signal cut-off of ≥8 million to determine antibody presence.^23,24^ Additional serology for IgG to nucleocapsid protein was performed using the Abbott Architect i2000 chemiluminescent microparticle immunoassay (Abbott, Maidenhead, UK). Antibody levels ≥1.40 arbitrary units were considered positive. PCR testing was performed on a range of platforms (see Supplementary Material).

### Statistical analysis

We classified HCWs according to their baseline antibody status. Those with only negative antibody tests, were considered at risk of infection from their first (negative) antibody test until the earlier of the study end (18-November-2020) or their first PCR-positive test. Those with a positive antibody test were considered at risk of (re)infection from 60 days after their first positive antibody result, to the earlier of study end or their next PCR-positive test, irrespective of subsequent sero-reversion, i.e. any later negative antibody test. The 60 day window was pre-specified to exclude PCR-positive tests due to RNA persistence from the index infection that led to seroconversion (based on earlier observations that RNA persistence could last for ≥6 weeks^25,26^). For the same reason, we also only considered PCR-positive tests occurring ≥60 days after the previous PCR-positive test. Those who were initially antibody-negative and then seroconverted were allowed to contribute to the analysis twice; once while at risk of infection and antibody-negative and then subsequently while antibody-positive.

We used Poisson regression to model incidence of PCR-positive infection per day at risk by baseline antibody status. Models adjusted for changes in overall incidence by including calendar month at risk as a categorical or calendar time as continuous variable (allowing for non-linear effects using natural cubic splines with up to 5 knots, at default positions, and choosing the final number of knots based on the best model fit using the Akaike information criterion). Additionally, we adjusted for age (allowing for non-linear effects similarly) and self-reported gender. We also fitted models considering baseline antibody titre instead of binary antibody status, using allowing for non-linear effects as above. Robust standard errors were used to account for some individuals contributing to the analysis twice, i.e. before and after seroconversion.

Primary analyses used anti-trimeric spike IgG assay results as these were expected a priori to relate most closely to neutralising activity and protection from infection.^7,10^ We also conducted two secondary analyses, considering anti-nucleocapsid antibodies and also a combined model where we allowed individuals to have one of three baseline antibody statuses (both assays negative, both positive, only one positive). Finally, we conducted a sensitivity analysis to investigate the impact of different asymptomatic testing rates by antibody status (see Supplementary Materials).

### Ethics statement

Deidentified data from staff testing were obtained from the Infections in Oxfordshire Research Database (IORD) which has generic Research Ethics Committee, Health Research Authority and Confidentiality Advisory Group approvals (19/SC/0403, 19/CAG/0144).

## Results

12219 HCWs had baseline anti-spike antibodies measured and were followed up between 23-April and 18-November-2020. 11052 (90.4%) HCWs were seronegative and 1167 (9.6%) were seropositive at their first anti-trimeric spike IgG measurement; 79 HCWs seroconverted during the study (Table 1, Figure S1). The median (IQR) [range] age was 38 (29-49) [16-86] years. Individuals were followed for a median (IQR) [range] 188 (174-195) [1-209] days after a negative antibody test and 127 (106-135) [1-149] days after a positive antibody test (starting the “at risk” period 60 days post positive-PCR/serology in seropositive individuals).

**Table 1.**
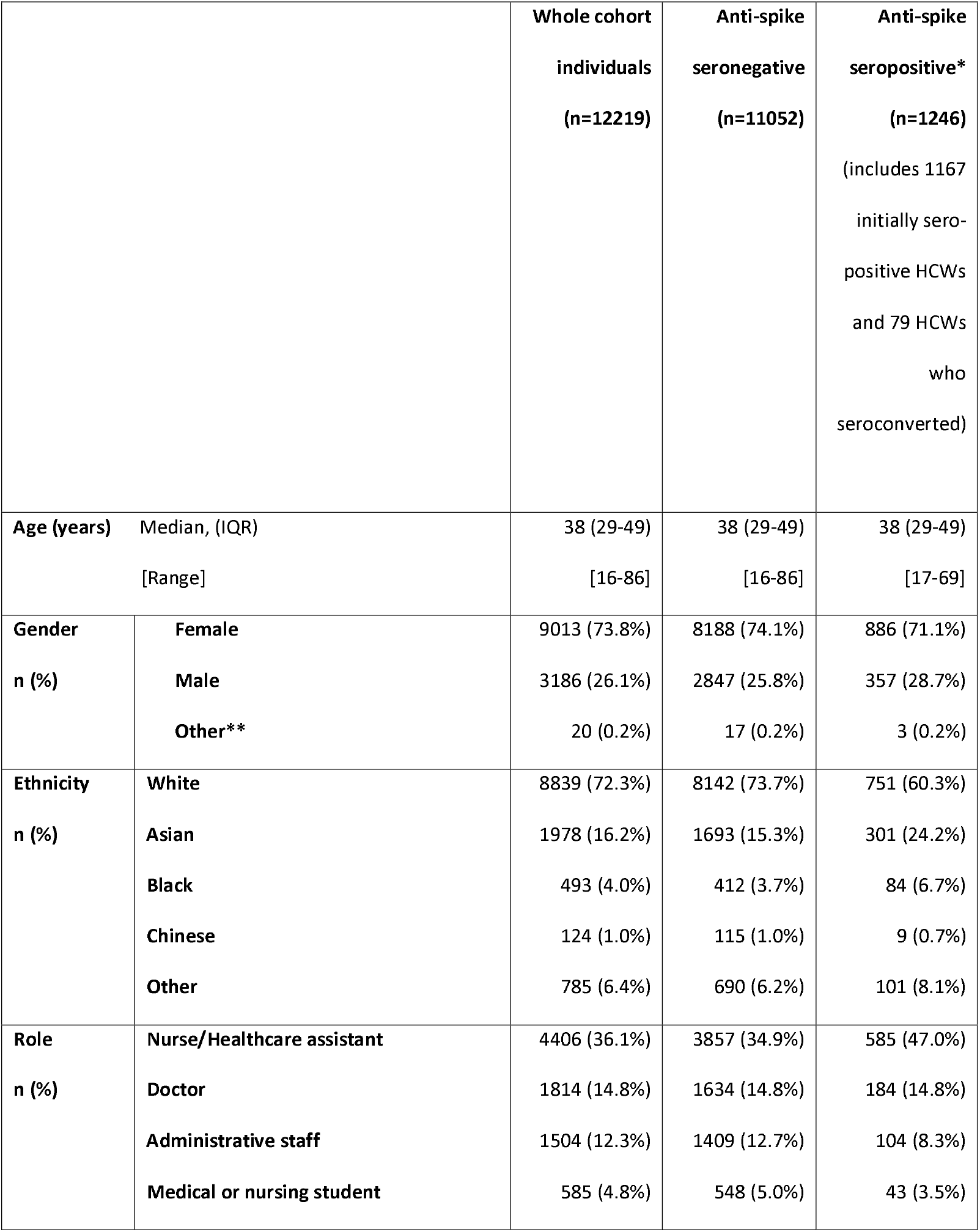
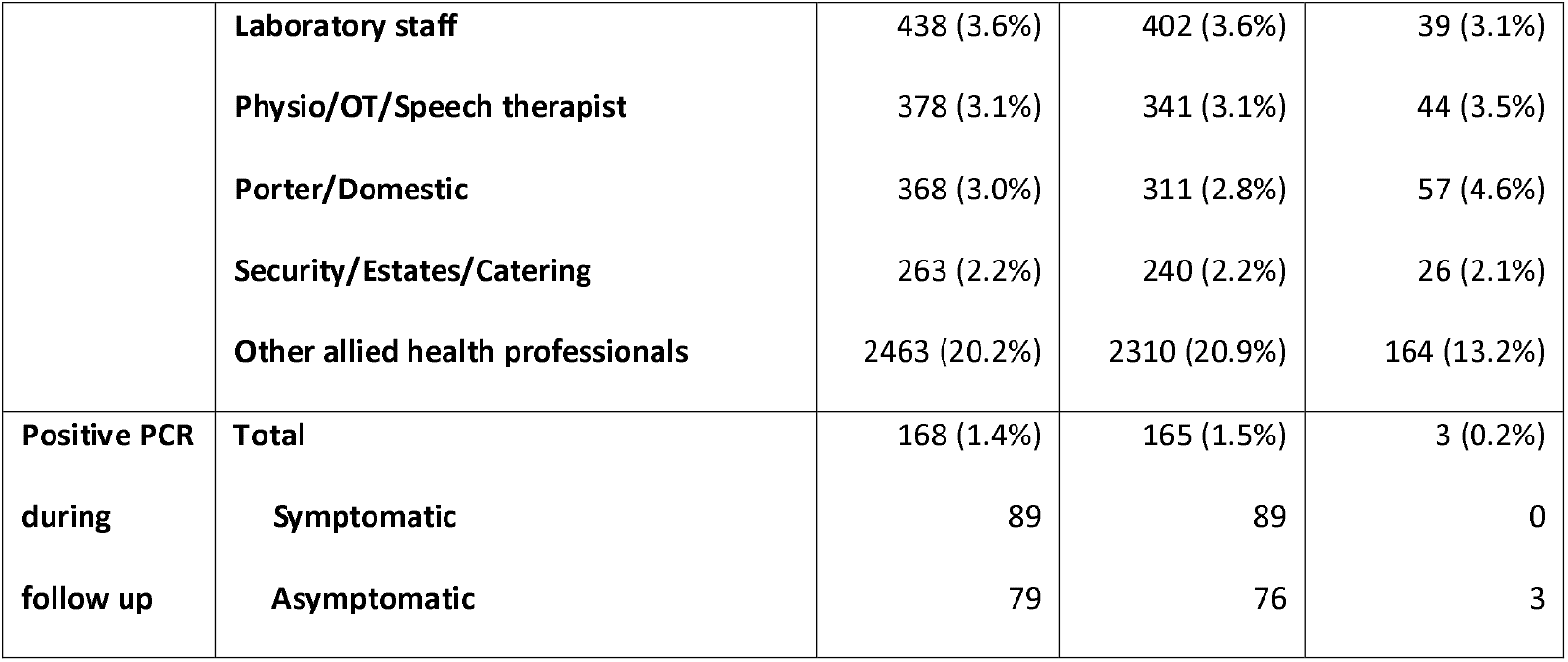
Demographics for 12219 healthcare workers with SARS-CoV-2 anti-spike IgG results. *Those who started anti-spike antibody negative and then seroconverted (n=79) were allowed to contribute to the analysis twice, once while at risk of infection and antibody negative and then subsequently while antibody positive and at risk of reinfection. Hence the sum of seropositive and seronegative subgroups is greater than the number of individuals in the whole cohort. **This category includes trans and non-disclosed gender, amalgamated due to small numbers to prevent individuals being identified.

Rates of symptomatic PCR testing were similar in seronegative and seropositive HCWs: 1597 PCR tests during 1,908,816 person-days of follow-up (8.4/10,000 person-days) and 106 during 143,022 person-days (7.4/10,000 person-days) respectively (rate ratio 0.90 [95%CI 0.74-1.09, p=0.27]). 8339 HCWs attended for ≥1 asymptomatic screen; seronegative HCWs attended for asymptomatic screening more frequently, (26,153 tests; 138/10,000 person-days), than seropositive staff (1442 tests; 102/10,000 person-days) (rate ratio 0.74 [95%CI 0.70-0.78, p<0.001]).

### Incidence of PCR positivity by baseline anti-spike IgG antibody status

165/11052 (1.5%) HCWs were PCR-positive while anti-spike IgG seronegative, 76 during asymptomatic screening and 89 while symptomatic. 3/1246 (0.2%) HCWs were PCR-positive following a positive anti-spike IgG measurement, all were asymptomatic. Allowing for the varying duration of follow-up, rates of new PCR-positive results were 0.86 and 0.21/10,000 days-at-risk in seronegative and seropositive HCWs respectively (incidence rate ratio, IRR, 0.24 [95%CI 0.08-0.76, p=0.015]). No antibody-positive individual had a subsequent symptomatic infection; rates of new PCR-confirmed symptomatic infection were 0.46 and 0.00/10,000 days-at-risk in seronegative and seropositive individuals respectively.

Incidence of PCR-positive results varied by calendar time (Figure 1A), reflecting the first and second waves of the pandemic, but was consistently higher in the seronegative versus seropositive group. Adjusting for age, gender and for changes in incidence by month, the IRR for being seropositive was 0.25 (95%CI 0.08-0.80, p=0.019) (Table 2) and adjusting for time as a continuous variable, 0.26 (95%CI 0.08-0.81, p=0.020) (Figure 1B). As rates of asymptomatic testing varied by antibody status, we performed a sensitivity analysis, randomly removing PCR results for seronegative HCWs to match testing rates in seropositive HCWs, yielding an adjusted IRR of 0.28 (95%CI 0.09-0.90, p=0.032).

**Table 2.** Estimated incidence rate ratios by anti-spike IgG antibody status as a binary variable adjusting for age, gender and incidence by calendar month. 20 HCWs identifying as Trans or with a non-disclosed gender are not shown, as there were zero PCR-positive results in these individuals, 17 of whom were seronegative and 3 of whom were seropositive. *Age was fitted as a continuous variable with a 5 knot spline (Supplementary Figure S2).

|  | Unadjusted<br>incidence<br>rate ratio | Unadjusted<br>95% CI | Unadjusted P<br>value | Adjusted<br>incidence<br>rate ratio | Adjusted<br>95% CI | Adjusted P<br>value |
| --- | --- | --- | --- | --- | --- | --- |
| Anti-spike<br>IgG negative | 1.00 |  |  | 1.00 |  |  |
| Anti-spike<br>IgG positive | 0.24 | 0.08, 0.76 | 0.015 | 0.25 | 0.08, 0.80 | 0.019 |
| April-June | 1.00 |  |  | 1.00 |  |  |
| July | 0.17 | 0.08, 0.37 | <0.001 | 0.18 | 0.08, 0.39 | <0.001 |
| August | 0.18 | 0.09, 0.38 | <0.001 | 0.20 | 0.09, 0.41 | <0.001 |
| September | 0.25 | 0.13, 0.48 | <0.001 | 0.27 | 0.14, 0.52 | <0.001 |
| October | 0.79 | 0.52, 1.19 | 0.26 | 0.86 | 0.57, 1.30 | 0.48 |
| November | 1.71 | 1.16, 2.51 | 0.006 | 1.87 | 1.27, 2.75 | 0.001 |
| Gender,<br>Female | 1.00 |  |  | 1.00 |  |  |
| Gender,<br>Male | 1.01 | 0.72, 1.43 | 0.93 | 1.03 | 0.73, 1.46 | 0.85 |
| Age* |  |  |  |  |  |  |

**Figure 1.**
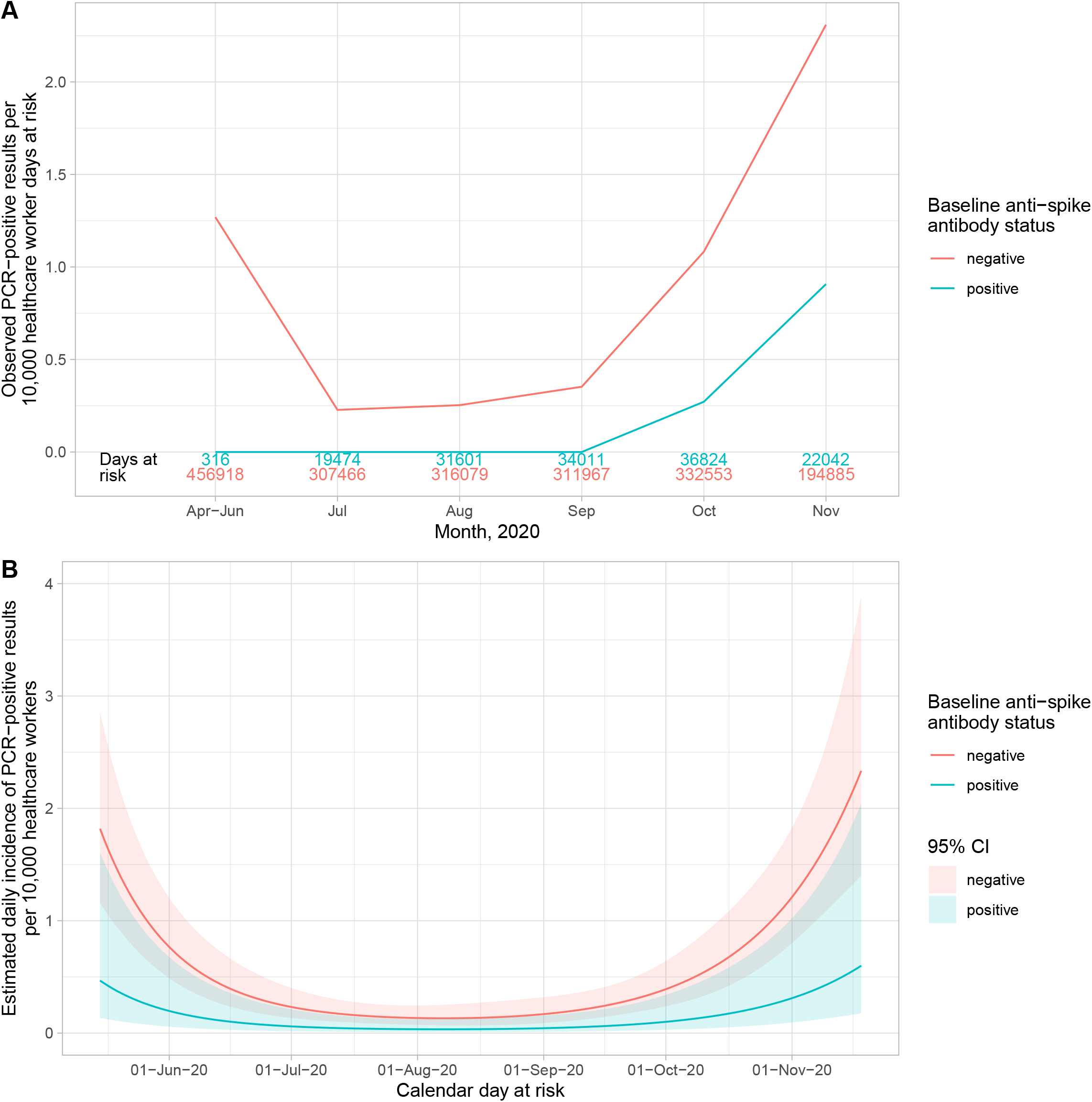
Observed and estimated incidence of SARS-CoV-2 positive PCR results by baseline anti-spike IgG antibody status. Panel A shows the observed cases per 10,000 HCW days at risk. The cases in seronegative staff are shown in red and seropositive staff in blue. The total number of HCW days at risk by month are shown in red and blue text above the x axis. Panel B shows the estimated daily incidence of SARS-CoV-2 positive PCR results per 10,000 HCW days at risk, by baseline antibody status (95% confidence intervals are indicated by the coloured ribbons). The Poisson regression model is adjusted for age (using a 5 knot spline, similar to Supplementary Figure S2), gender and calendar time fitted as continuous, using a 5 knot natural cubic spline with default knot positions.

### Incidence of PCR positivity by baseline antibody titre

In addition to the categorical antibody result, we analysed numerical anti-spike IgG titres. Adjusting for month of testing, age and gender, the estimated incidence of PCR-positive results started to fall from titres of ∼4 million and above (cf. threshold for “detection” of 8 million^23^) (Figure 2A).

**Figure 2.**
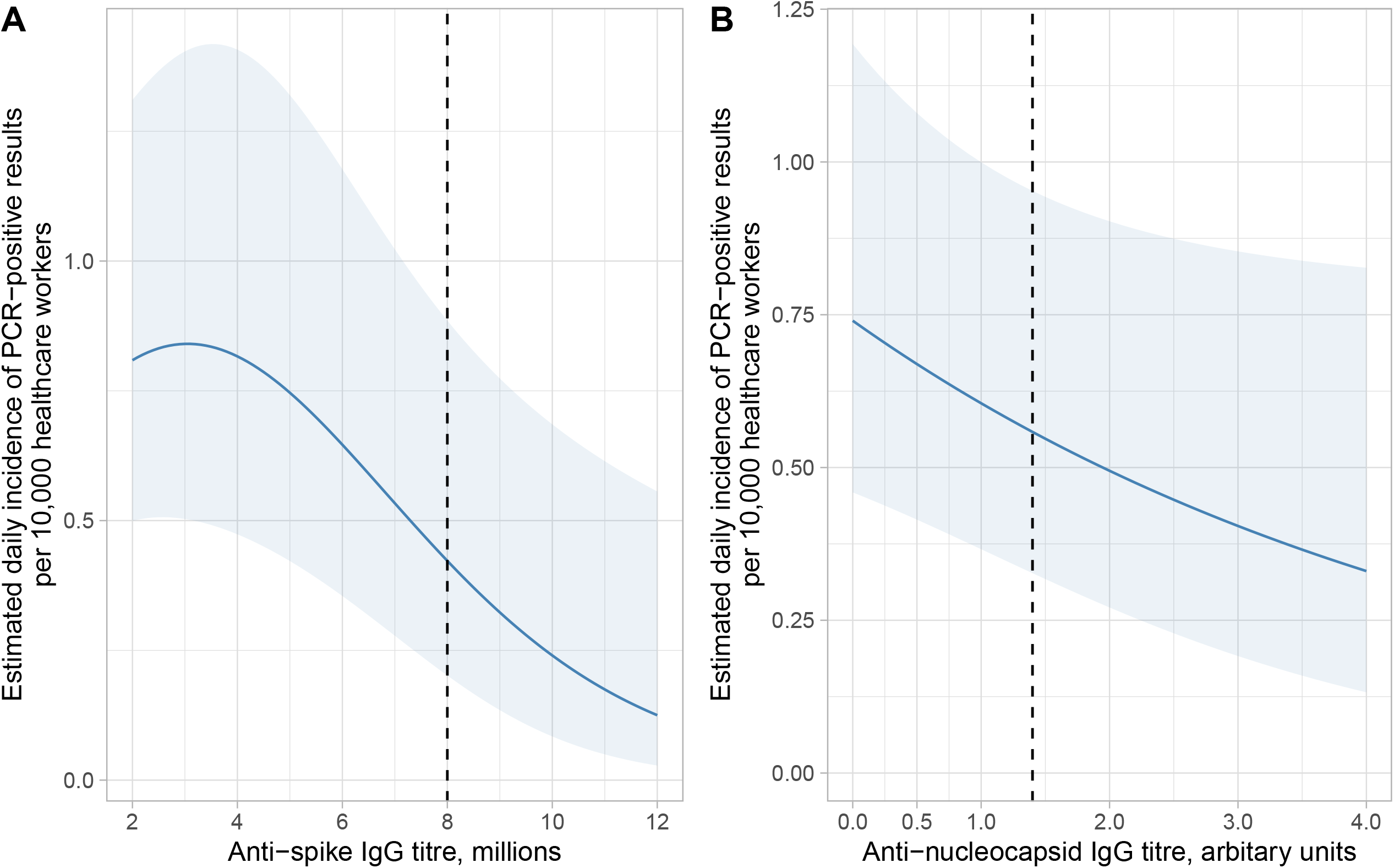
Estimated daily incidence by baseline anti-trimeric spike and anti-nucleocapsid spike IgG antibody titre. Panel A shows the estimated daily incidence per 10,000 healthcare workers, by anti-trimeric spike IgG titre (Oxford immunoassay, p-value vs no trend = 0.013), using a 3 knot spline (number of knots chosen using the Akaike information criterion [AIC]). Panel B shows the estimated daily incidence per 10,000 healthcare workers by anti-nucleocapsid IgG titre (Abbott SARS-CoV-2 IgG assay; p=0.08), linear model plotted as lowest AIC. The models used in both panels adjust for age (set at the median age, 38 years), gender (set as female) and calendar month (set as October 2020). The light blue ribbon shows the 95% confidence interval, and the vertical dotted line indicates the assay positive cut-off.

### Secondary analyses using anti-nucleocapsid IgG

We conducted two secondary analyses. Firstly, using anti-nucleocapsid instead of anti-spike antibody measurements as a marker for prior infection (n=12453 HCWs; Figure S1 and Table S1). 169/11336 (0.88/10,000 person-days) seronegative HCWs tested PCR-positive compared to 3/1162 (0.21/10,000 person-days) antibody-positive HCWs. Accounting for time at risk, age, gender and calendar month, the adjusted IRR for those antibody-positive was 0.26 (95%CI 0.08-0.81, p=0.021, Table S2). There was marginal evidence that the incidence of PCR-positive results fell with increasing anti-nucleocapsid antibody titres (IRR per unit increase 0.82 [95%CI 0.65-1.03, p=0.08], Figure 2B).

Second, we classified the 12153 HCWs with available anti-spike and anti-nucleocapsid baseline results, as both assays negative, both positive, or only one positive (Figure S1, Tables S3 and S4). 160/10866 (0.85/10,000 person-days) of those with two negative assays had subsequent PCR-positive tests, versus 2/1015 (0.16/10,000 person-days) with both baseline assays positive and 2/333 (0.55/10,000 person-days) with mixed antibody results. There were reduced rates of subsequent PCR-positive tests in those where both antibody tests were positive compared to both negative, adjusting for age, gender and calendar month, IRR 0.20 (95%CI 0.05-0.81, p=0.024), but an absence of evidence in those with mixed results, adjusted IRR 0.67 (0.16-2.71, p=0.57).

### Description of seropositive individuals with subsequent PCR-positive results

Four seropositive HCWs subsequently tested PCR-positive (three with anti-spike IgG, two of whom had anti-nucleocapsid IgG, and one additional with anti-nucleocapsid IgG only) (Figure 3 and Table S5).

**Figure 3.**
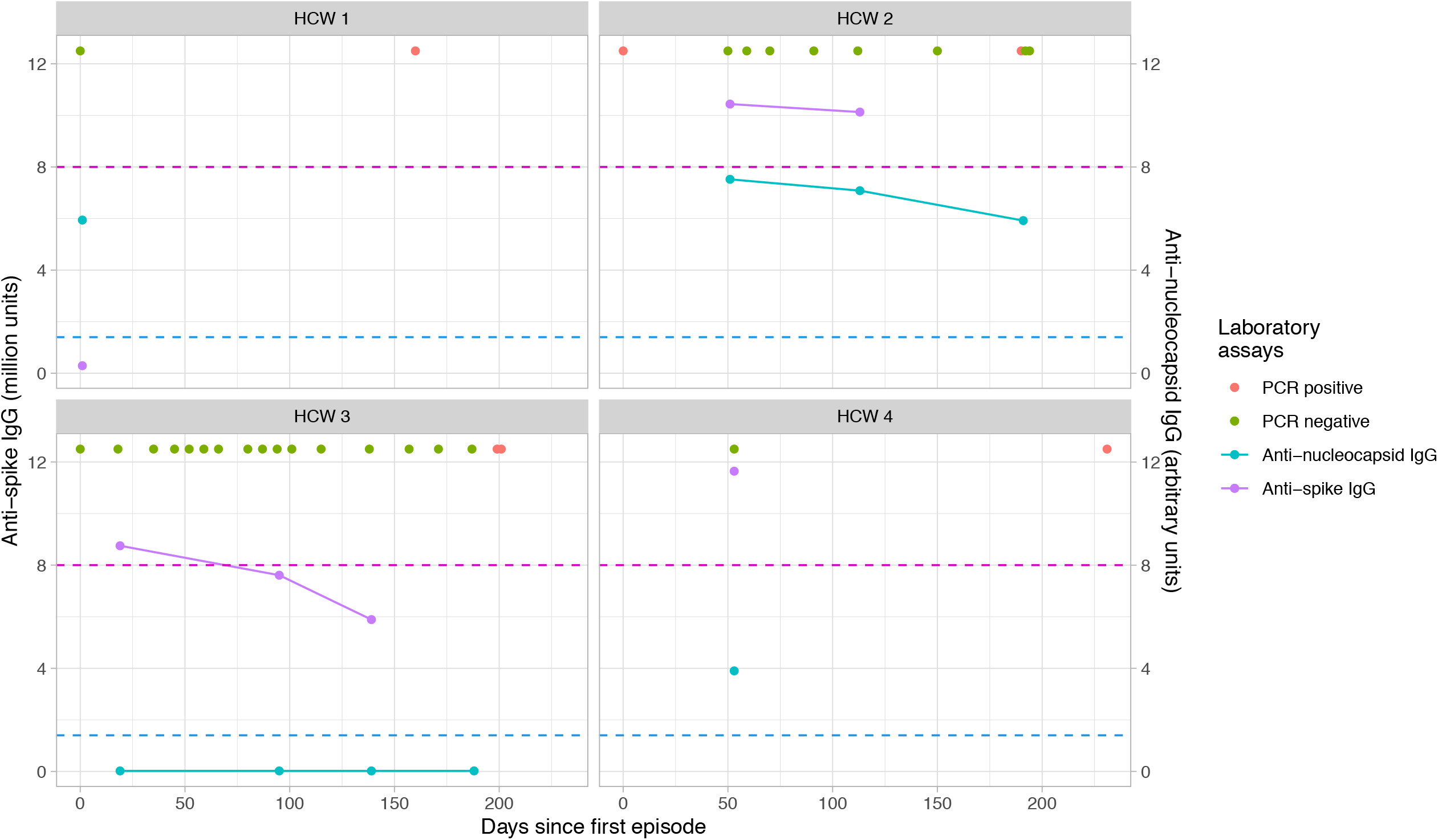
Antibody trajectories and PCR results for four seropositive individuals with subsequent positive PCR results. Three HCWs became PCR-positive having been anti-spike IgG seropositive (two also positive for anti-nucleocapsid IgG), and one having been anti-nucleocapsid IgG positive alone. The x axis shows time since the first episode, this is defined as date of first positive PCR, onset of symptoms if symptomatic and no PCR performed, or first attendance at clinic if asymptomatic and no PCR performed during presumed index infection. Antibody trajectories are shown by lines with circular points (blue = anti-nucleocapsid IgG, lilac = anti-spike IgG), PCR results are shown in green (negative) and red (positive). Assay thresholds are shown by dotted lines; anti-spike IgG (positive ≥8 million (lilac dotted line)) assays and anti-nucleocapsid IgG (positive ≥1.4 (blue dotted line).

HCW1 was seropositive for anti-nucleocapsid IgG when first tested in May (original sample retrieved and re-tested, confirmed positive), but seronegative for anti-spike IgG. They had no history of prior Covid-19-like symptoms and no prior positive PCR. They developed a febrile illness 160 days later confirmed as Covid-19 by PCR.

HCW2 presented with Covid-19-like symptoms in April and was PCR-positive; they seroconverted with positive anti-spike and anti-nucleocapsid antibodies and remained antibody positive on both assays up to October. They had 5 negative PCRs before becoming PCR-positive again at day 190, diagnosed through asymptomatic staff testing and could not recall any symptoms in retrospect. Repeat PCR tests performed 2 and 4 days later were negative.

HCW3 reported a febrile illness in mid-February and was not tested for SARS-CoV-2 at the time. They were seropositive for anti-spike, but not anti-nucleocapsid antibodies in May. Three subsequent antibody tests between July and October were negative for both antibodies, with evidence of waning anti-spike titres and persistent low anti-nucleocapsid titres (Figure 3). They regularly attended asymptomatic screening, and had 14 negative PCRs, becoming PCR-positive 180 days following their first serological test. They were asymptomatic when tested, but reported transient myalgia following an influenza vaccine the previous week. Repeat PCR 2 days later was positive.

HCW 4 developed fever and anosmia in March but did not have PCR test. They were seropositive for anti-spike and anti-nucleocapsid antibodies when first tested in May. They tested PCR-positive, whilst asymptomatic, 231 days following initial symptoms.

## Discussion

Virus-specific antibodies are usually recognised as correlates or surrogates of antiviral immunity. Here we describe the results of over 2 million person-days of follow-up, from 11052 HCWs following negative serology, and 1246 following a positive anti-spike antibody test. We show in our primary analysis, that evidence of previous SARS-CoV-2 infection, defined by presence of anti-spike antibodies, is associated with lower risk of a repeat PCR-positive test (IRR 0.24 [95%CI 0.08-0.76, p=0.015]). No symptomatic infections and only three asymptomatic PCR-positive results were seen in those with anti-spike antibodies, over 30 weeks of follow-up. This suggests that antibodies produced by prior SARS-CoV-2 infection are associated with protection from reinfection for most people for at least six months. Evidence of post-infection immunity was also seen when anti-nucleocapsid IgG, or the combination of both anti-nucleocapsid and anti-spike IgG were used as markers of prior infection.

The incidence of SARS-CoV-2 infection was inversely associated with baseline anti-spike antibody titres, with higher antibody titres associated with the lowest incidence of positive PCR results. This association extended below the positive threshold for the assay (8 million), with likelihood of infection falling above 4 million units. It is likely that some of the individuals with baseline titres below the assay cut-off were previously infected with SARS-CoV-2, and either had low peak antibody titres post infection, or may have been tested later post-infection when antibody titres had declined below the assay threshold.^5^ There was also marginal evidence that rates of PCR-positive results went down with higher anti-nucleocapsid IgG levels.

Notably two of the four seropositive individuals with subsequent PCR-positive tests had discordant baseline antibody results which were positive on only one of the two IgG assays, with low negative readings on the other assay. Neither had a PCR-confirmed primary infection. It is plausible that one or both had false-positive antibody results which may have occurred for a number of reasons including immunoassay interference.^27^ One of the two developed subsequent symptomatic infection. The two other HCWs seroconverted with both anti-spike and anti-nucleocapsid antibodies detected after an initial illness, one PCR confirmed (albeit with a borderline Ct value). One had a single PCR-positive test, and the other a transient subsequent PCR-positive result, not detected on repeat testing. Both results are consistent with SARS-CoV-2 re-exposure that did not lead to repeat symptoms.

Due to the low number of reinfections in the baseline seropositive group, this study is not currently powered to detect if past seroconversion or current antibody levels are more strongly associated with protection from infection. Similarly, we cannot say how past infection confers protection, whether specifically through the antibodies we measured, or T cell immunity which we did not. Our study is relatively short, following HCWs up for 30 weeks. Ongoing longitudinal measurements of markers of humoral and cellular immunity, and PCR testing, will be required to determine the duration and determinants of protection.

HCWs were enrolled in a voluntary testing scheme, with a flexible follow-up schedule, leading to different attendance frequencies. Although asymptomatic PCR testing was offered up to every 2 weeks, HCWs attended less frequently (mean: once every 10-14 weeks), in particular when seropositive. Therefore, asymptomatic infection is likely to have been under-ascertained in both groups. Additionally, as staff were told their antibody results, this led to ‘outcome ascertainment bias’, with staff attending less frequently if seropositive. However, a sensitivity analysis suggests the differing attendance rates did not substantially alter our findings. Staff were told explicitly to continue to practice social distancing and follow personal protective equipment guidance irrespective of their antibody results and to attend for testing if they developed fever, new cough or anosmia/ageusia even if they had previously tested PCR or antibody-positive. This is reflected in the similar rates of attendance for symptomatic testing observed if seropositive or seronegative.

Some staff have been lost to follow-up due to terminating employment at our hospitals; however this is likely to have occurred at similar rates in symptomatic and asymptomatic staff. Some PCR-positive results from government symptomatic testing sites may not have been notified to the hospital. Finally, as this is a study of predominantly working age HCWs, further studies are needed to assess post-infection immunity in children, the over 60s and those with comorbidities, particularly the immunosuppressed.

In summary, this study suggests that prior SARS-CoV-2 infection (defined by either anti-spike or anti-nucleocapsid antibodies) offers protection from reinfection, in the short term. However, lack of knowledge regarding the specific effects of each of the range of antibody assays in use, the precise extent of protection and long-term immune responses do not support exempting those positive for anti-SARS-CoV-2 antibodies from infection control and public health pandemic control measures at present. Further longitudinal studies, over longer durations, looking at the full range of immune responses to SARS-CoV-2, will be required to determine the full range of correlates and surrogates of protective immunity, the duration of post-infection immunity to reinfection, disease, hospitalisation or death and its impact on onward transmission. This will have important implications for public policy, guiding behaviour and infection control in healthcare settings and beyond.

## Data Availability

The data studied are available from the Infections in Oxfordshire Research Database (https://oxfordbrc.nihr.ac.uk/research-themes-overview/antimicrobial-resistance-and-modernising-microbiology/infections-in-oxfordshire-research-database-iord/), subject to an application and research proposal meeting the ethical and governance requirements of the Database. For further details on how to apply for access to the data and for a research proposal template please.

## Acknowledgements

We would like to thank all OUH staff who participated in the staff testing programme, and the staff and medical students who ran the programme. This work uses data provided by healthcare workers and collected by the UK’s National Health Service as part of their care and support. We thank all the people of Oxfordshire who contribute to the Infections in Oxfordshire Research Database. Research Database Team: L Butcher, H Boseley, C Crichton, DW Crook, DW Eyre, O Freeman, J Gearing (community), R Harrington, K Jeffery, M Landray, A Pal, TEA Peto, TP Quan, J Robinson (community), J Sellors, B Shine, AS Walker, D Waller. Patient and Public Panel: G Blower, C Mancey, P McLoughlin, B Nichols.

