## Supplementary Material for "Antibodies to SARS-CoV-2 are associated with protection against reinfection"

### Supplementary Methods

#### PCR platforms

RT-PCR was performed using the Public Health England SARS-CoV-2 assay (targeting the RdRp gene), one of five commercial assays: Abbott RealTime (targeting RdRp and N genes; Abbott, Maidenhead, UK), Altona RealStar (targeting E and S genes; Altona Diagnostics, Liverpool, UK), Cepheid Xpert® Xpress SARS-CoV-2 (targeting N2 and E; Cepheid, California, USA), BioFire® Respiratory 2.1 (RP2.1) panel with SARS-CoV-2 (targeting ORF1ab and ORF8; Biofire diagnostics, Utah, USA), Thermo Fisher TaqPath assay (targeting S and N genes, and ORF1ab; Thermo Fisher, Abingdon, UK) or using the ABI 7500 platform (Thermo Fisher, Abingdon, UK) with the US Centers for Disease Control and Prevention Diagnostic Panel of two probes targeting the N gene.

#### Software

All analyses were performed using R, version 3.6.3. Natural cubic splines were fitted with the splines library, using the default spline locations.

#### Sensitivity analysis

Rates of asymptomatic testing varied by antibody status, with seronegative healthcare workers (HCWs) attending more frequently than seropositive HCWs. To assess the impact on our results we performed a sensitivity analysis where we randomly removed PCR tests from the dataset. PCR tests were removed at random, irrespective of the result, from individuals with negative baseline serology until the overall rate of testing per 10,000 days at risk was equivalent in the seropositive and seronegative HCW groups. We used the resulting dataset to repeat our analysis.

### Supplementary Figures

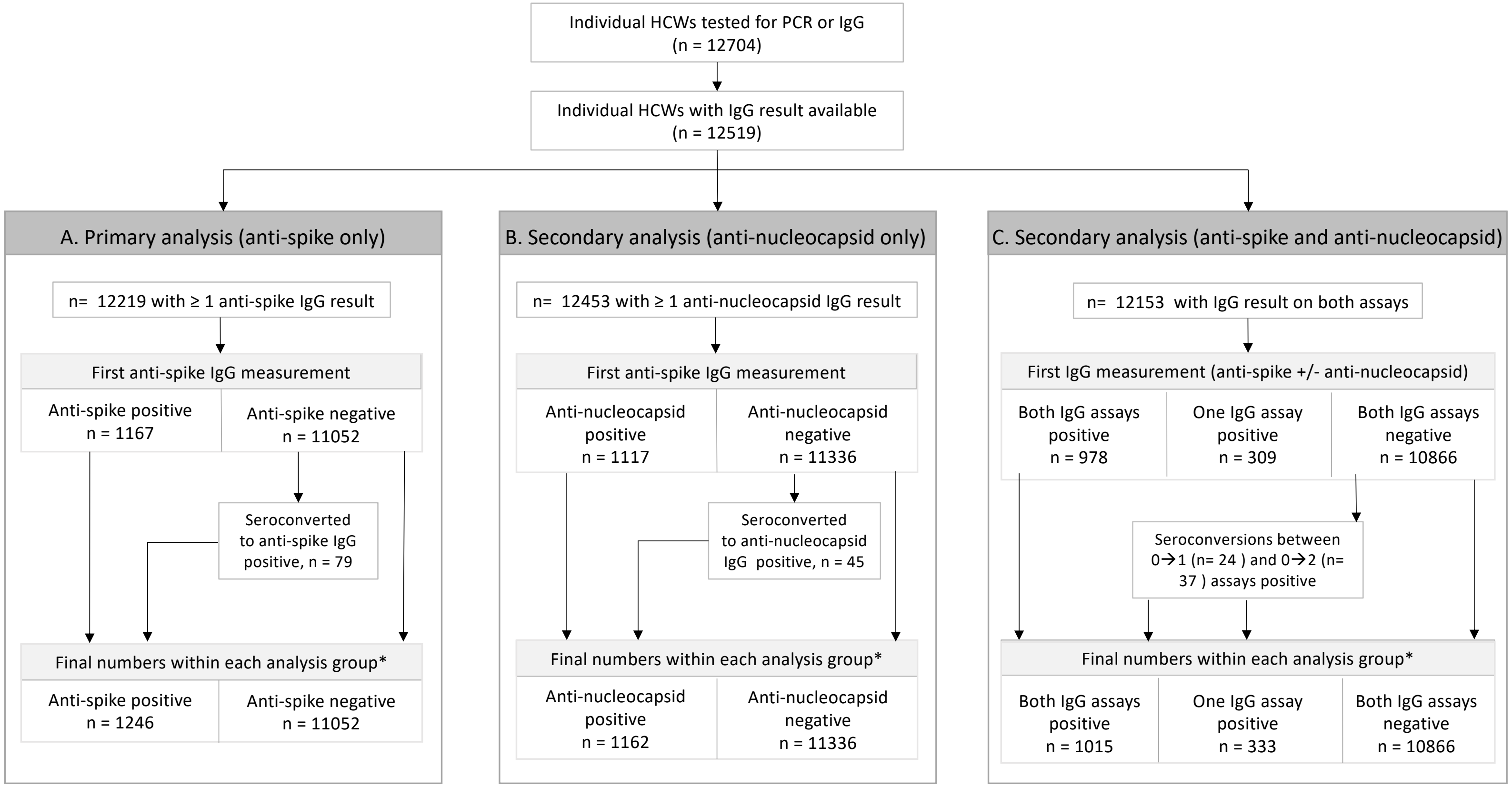


* NB HCWs who seroconvert during follow up were allowed to contribute to the analysis twice, once while at risk of infection and IgG negative and then subsequently while IgG positive.

**Supplementary Figure S1. Flow diagram demonstrating cohort numbers from enrollment to final categorization into seropositive and seronegative groups.** Panel A shows the cohort used in the primary analysis, using anti-spike IgG results only. Panel B shows the cohort used in the first secondary analysis using anti-nucleocapsid IgG results only. Panel C shows the cohort used in the second secondary analysis, using both anti-spike and anti-nucleocapsid IgG results.

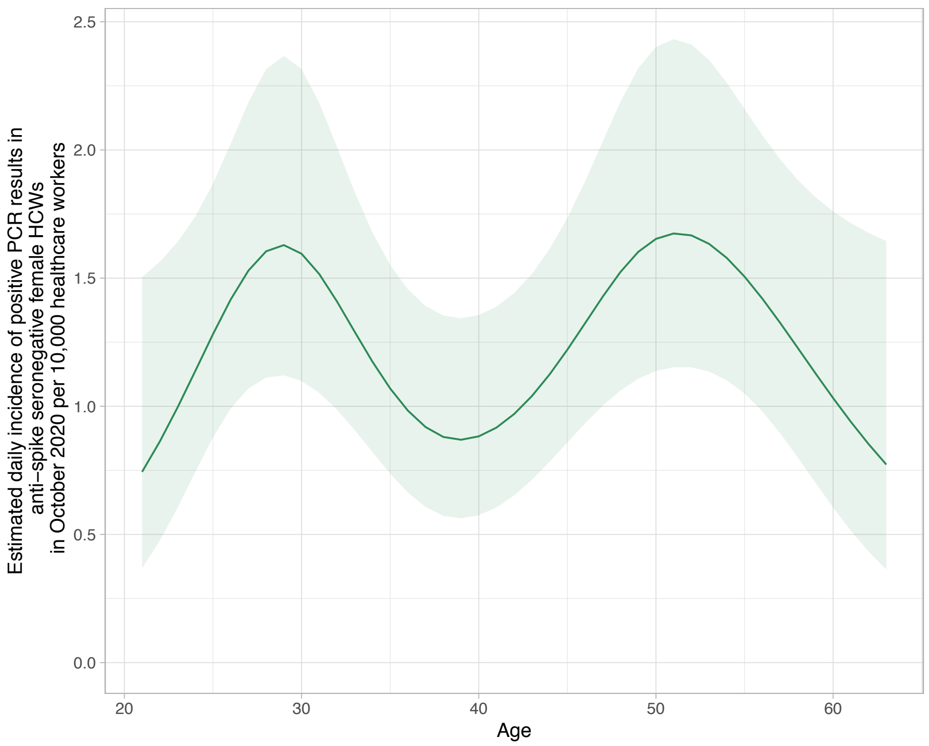


**Supplementary Figure S2. Non-linear relationship between age and incidence of PCR-positive results in HCWs with anti-spike antibody measurements**. Estimates shown are adjusted for gender, month, and baseline antibody status.

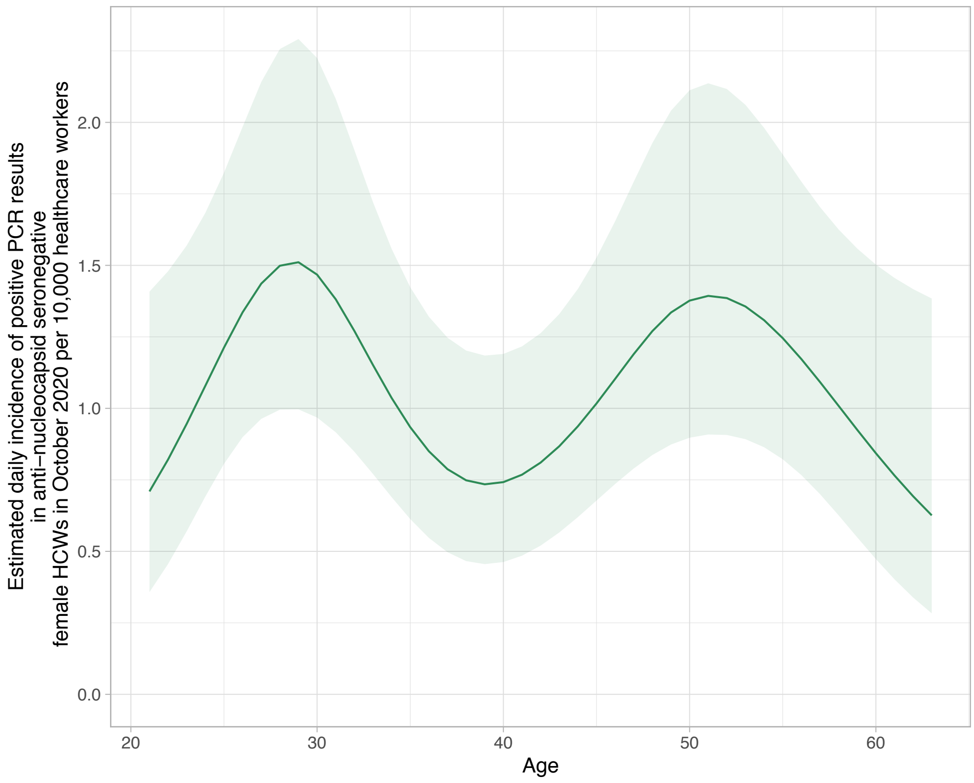


**Supplementary Figure S3. Non-linear relationship between age and incidence of PCR-positive results in HCWs with anti-nucleocapsid antibody measurements**. Estimates shown are adjusted for gender, month, and baseline antibody status.

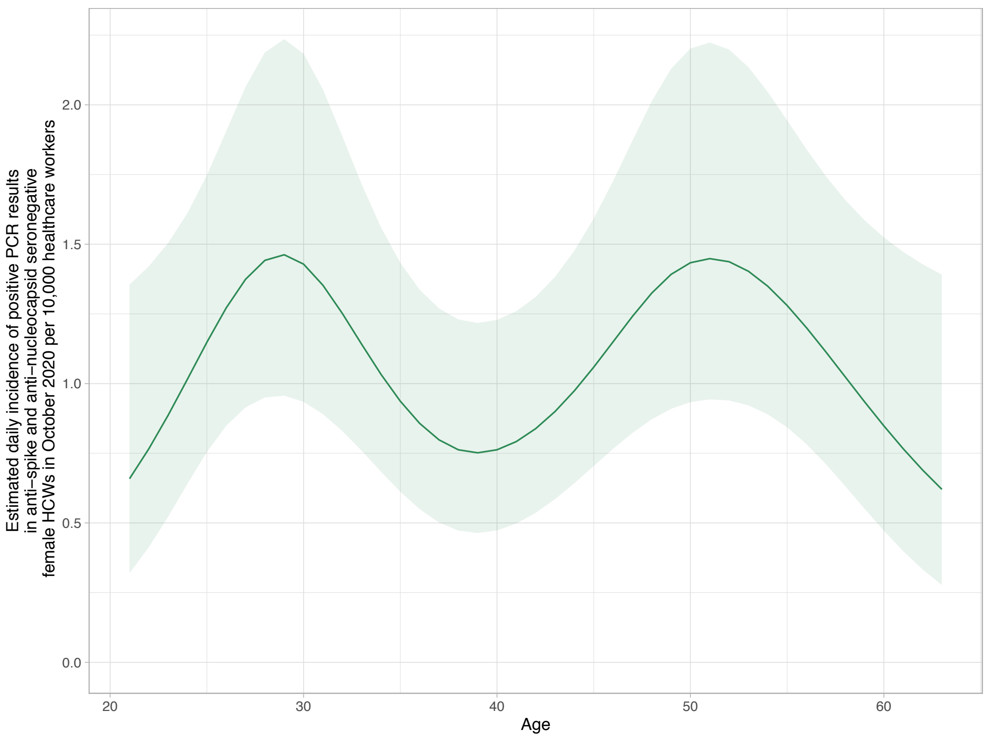


**Supplementary Figure S4. Non-linear relationship between age and incidence of PCR-positive results in HCWs with both anti-spike and anti-nucleocapsid antibodies**. Estimates shown are adjusted for gender, month, and baseline antibody status.

### Supplementary Tables

| | | Whole cohort (n=12453) | First secondary analysis (anti-nucleocapsid IgG only) | |
|---|---|---|---|---|
| | | | **Anti-nucleocapsid seronegative (n=11336)** | **Anti-nucleocapsid seropositive** (includes 1117 initially seropositive HCWs and 45 who seroconverted)* **(n=1162)** |
| **Age (years)** Median (IQR), [Range] | | 38 (29-49) [16-86] | 38 (28-49) [16-86] | 39 (29-49) [17-70] |
| **Gender n (%)** | Female | 9187 (73.8%) | 8399 (74.1%) | 822 (70.7%) |
| | Male | 3246 (26.1%) | 2919 (25.7%) | 338 (29.1%) |
| | Other** | 20 (0.2%) | 18 (0.2%) | 2 (0.2%) |
| **Ethnicity n (%)** | White | 8992 (72.2%) | 8336 (73.5%) | 685 (59) |
| | Asian | 2019 (16.2%) | 1737 (15.3%) | 290 (25%) |
| | Black | 505 (4.1%) | 427 (3.8%) | 78 (6.7%) |
| | Chinese | 130 (1.0%) | 121 (1.1%) | 9 (0.8%) |
| | Other | 807 (6.5%) | 715 (6.3%) | 100 (8.6%) |
| **Role n (%)** | Nurse/HCA | 4471 (35.9%) | 3929 (34.7%) | 562 48.4% |
| | Doctor | 1847 (14.8%) | 1674 (14.8%) | 175 15.1% |
| | Administrative staff | 1527 (12.3%) | 1445 (12.7%) | 89 7.7% |
| | Medical or nursing student | 632 (5.1%) | 606 (5.3%) | 30 2.6% |
| | Laboratory staff | 443 (3.6%) | 409 (3.6%) | 35 3% |
| | Physio/OT/Speech therapist | 385 (3.1%) | 346 (3.1%) | 41 3.5% |
| | Porter/Domestic | 376 (3%) | 317 (2.8%) | 59 5.1% |
| | Security/Estates/Catering | 268 (2.2%) | 242 (2.1%) | 30 2.6% |
| | Other allied health professionals | 2504 (20.1%) | 2368 (20.9%) | 141 12.1% |
| **PCR-positive during follow up (n)** | Total | 172 | 169 | 3 |
| | Symptomatic | 89 | 88 | 1 |
| | Asymptomatic | 83 | 81 | 2 |

**Supplementary Table S1. Baseline cohort demographics for 12453 healthcare workers included in the secondary analysis using anti-nucleocapsid IgG alone.**
*Those who started anti-nucleocapsid antibody negative and then seroconverted (n=45) were allowed to contribute to the analysis twice, once while at risk of infection and antibody negative and then subsequently while antibody positive and at risk of re-infection.
**This category includes Trans and non-disclosed gender, amalgamated due to small numbers to prevent inadvertent identification.

| | Unadjusted incidence rate ratio | Unadjusted 95% CI | Unadjusted P value | Adjusted incidence rate ratio | Adjusted 95% CI | Adjusted P value |
|---|---|---|---|---|---|---|
| **Anti-spike IgG negative** | 1.00 | | | 1.00 | | |
| **Anti-spike IgG positive** | 0.24 | 0.08, 0.76 | 0.015 | 0.26 | 0.08, 0.81 | 0.021 |
| **April-June** | 1.00 | | | 1.00 | | |
| **July** | 0.17 | 0.08, 0.37 | <0.001 | 0.18 | 0.08, 0.39 | <0.001 |
| **August** | 0.18 | 0.09, 0.38 | <0.001 | 0.19 | 0.09, 0.41 | <0.001 |
| **September** | 0.27 | 0.15, 0.51 | <0.001 | 0.29 | 0.16, 0.55 | <0.001 |
| **October** | 0.78 | 0.52, 1.18 | 0.25 | 0.85 | 0.56, 1.28 | 0.44 |
| **November** | 1.78 | 1.22, 2.60 | 0.003 | 1.94 | 1.32, 2.83 | 0.001 |
| **Gender, Female** | 1.00 | | | 1.00 | | |
| **Gender, Male** | 0.93 | 0.65,1.31 | 0.66 | 0.94 | 0.66, 1.33 | 0.73 |
| **Age*** | | | | | | |

**Supplementary Table S2. Estimated regression parameters for the Poisson regression with anti-nucleocapsid antibody status as a binary variable adjusting for calendar month.** 20 HCWs identifying as Trans or with a non-disclosed gender are not shown, as there were zero PCR-positive results in these individuals, 18 of whom were seronegative and 2 of whom were seropositive. *Age was fitted as a continuous variable with a 5 knot spline (Supplementary Figure S3).

| | | Whole cohort (n=12153) | Second secondary analysis (anti-spike +/- anti-nucleocapsid IgG) | | | |
|---|---|---|---|---|---|---|
| | | | Both assays negative (n=10866) | Anti-nucleocapsid positive only (n=135) | Anti-spike positive only (n=198) | Both assays positive* (n=1015) |
| **Age**<br>Median (IQR), [Range] | | 38 (29-49) [16-86] | 38 (29-49) [16-86] | 35 (27-47) [20-70] | 34 (26-44) [19-67] | 40 (30-49) [17-69] |
| **Gender**<br>**n (%)** | Female | 8970 (73.8%) | 8053 (74.1%) | 101 (74.8%) | 147 (74.2%) | 713 (70.2%) |
| | Male | 3163 (26.0%) | 2796 (25.7%) | 34 (25.2%) | 50 (25.3%) | 300 (29.6%) |
| | Other** | 20 (0.2%) | 17 (0.2%) | 0 (0.0%) | 1 (0.5%) | 2 (0.2%) |
| **Ethnicity**<br>**n (%)** | White | 8792 (72.3%) | 8012 (73.7%) | 92 (68.1%) | 141 (71.2%) | 588 (57.9%) |
| | Asian | 1963 (16.2%) | 1656 (15.2%) | 27 (20.0%) | 32 (16.2%) | 261 (25.7%) |
| | Black | 490 (4.0%) | 404 (3.7%) | 6 (4.4%) | 10 (5.1%) | 71 (7.0%) |
| | Chinese | 124 (1.0%) | 114 (1.0%) | 1 (0.7%) | 2 (1.0%) | 7 (0.7%) |
| | Other | 784 (6.5%) | 680 (6.3%) | 9 (6.7%) | 13 (6.6%) | 88 (8.7%) |
| **Role**<br>**n (%)** | Nurse/HCA | 4390 (36.1%) | 3788 (34.9%) | 58 (43.0%) | 71 (35.9%) | 499 (49.2%) |
| | Doctor | 1797 (14.8%) | 1593 (14.7%) | 26 (19.3%) | 35 (17.7%) | 147 (14.5%) |
| | Administrative staff | 1496 (12.3%) | 1392 (12.8%) | 10 (7.4%) | 23 (11.6%) | 79 (7.8%) |
| | Medical or nursing student | 584 (4.8%) | 545 (5.0%) | 2 (1.5%) | 16 (8.1%) | 26 (2.6%) |
| | Laboratory staff | 432 (3.6%) | 392 (3.6%) | 4 (3.0%) | 7 (3.5%) | 31 (3.1%) |
| | Physio/OT/Speech therapist | 377 (3.1%) | 331 (3.0%) | 11 (8.1%) | 8 (4.0%) | 30 (3.0%) |
| | Porter/Domestic | 367 (3.0%) | 304 (2.8%) | 7 (5.2%) | 5 (2.5%) | 51 (5.0%) |
| | Security/Estates/Catering | 260 (2.1%) | 234 (2.2%) | 4 (3.0%) | 1 (0.5%) | 25 (2.5%) |
| | Other allied health professionals | 2450 (20.2%) | 2287 (21.0%) | 13 (9.6%) | 32 (16.2%) | 127 (12.5%) |
| **Positive PCR during follow up** | Total | | 160 | 1 | 1 | 2 |
| | Symptomatic | | 87 | 1 | 0 | 0 |
| | Asymptomatic | | 73 | 0 | 1 | 2 |

**Supplementary Table S3. Baseline cohort demographics for 12153 healthcare workers included in the secondary analysis using a combination of anti-nucleocapsid and anti-spike IgG.** *Those who started antibody negative and then seroconverted were allowed to contribute to the analysis twice, once while at risk of infection and antibody negative and then subsequently while antibody positive and at risk of re-infection. **This category includes Trans and non-disclosed gender, amalgamated due to small numbers to prevent inadvertent identification.

| | Unadjusted incidence rate ratio | Unadjusted 95% CI | Unadjusted P value | Adjusted incidence rate ratio | Adjusted 95% CI | Adjusted P value |
|---|---|---|---|---|---|---|
| **Both IgG assays negative** | 1.00 | | | 1.00 | | |
| **Both IgG assays positive** | 0.19 | 0.05, 0.78 | 0.021 | 0.20 | 0.05, 0.81 | 0.024 |
| **Only one IgG assay positive** | 0.65 | 0.16, 2.62 | 0.54 | 0.67 | 0.16, 2.71 | 0.57 |
| **April-June** | 1.00 | | | 1.00 | | |
| **July** | 0.18 | 0.08, 0.39 | <0.001 | 0.19 | 0.09, 0.42 | <0.001 |
| **August** | 0.19 | 0.09, 0.40 | <0.001 | 0.21 | 0.10, 0.44 | <0.001 |
| **September** | 0.27 | 0.14, 0.51 | <0.001 | 0.29 | 0.15, 0.55 | <0.001 |
| **October** | 0.84 | 0.55, 1.27 | 0.40 | 0.91 | 0.60, 1.38 | 0.67 |
| **November** | 1.81 | 1.22, 2.67 | 0.003 | 1.98 | 1.33, 2.93 | 0.001 |
| **Gender, Female** | 1.00 | | | 1.00 | | |
| **Gender, Male** | 0.99 | 0.69, 1.40 | 0.94 | 1.00 | 0.71, 1.43 | 0.99 |
| **Age*** | | | | | | |

**Supplementary Table S4. Estimated regression parameters for the Poisson regression with both anti-spike and anti-nucleocapsid IgG antibody status adjusting for calendar month.** 20 HCWs identifying as Trans or with a non-disclosed gender are not shown, as there were zero PCR-positive results in these individuals, 17 of whom were seronegative on both tests and 2 of whom were seropositive on both tests and 1 who was seropositive on only one test. *Age was fitted as a continuous variable with a 5 knot spline (Supplementary Figure S4). Of 333 tests where only one assay was positive, 135 were positive for anti-nucleocapsid only and 198 for anti-spike only.

| ID | Demo-graphics | Baseline serology | Time between episodes* | Clinical characteristics | Timing of PCR, Ct value and assay |
|---|---|---|---|---|---|
| HCW 1 | 28F, White | Anti-spike: not detected (<1 million)<br><br>**Anti-nucleocapsid: detected (5.94)** | 160 days | 1st episode: asymptomatic<br><br>2nd episode: symptomatic (mild, febrile illness) | 1st episode: not done (prior to start of asymptomatic testing)<br><br>2nd episode: Cn 10.6 (Abbott, RdRp/N assay), not repeated |
| HCW 2 | 56F, White | **Anti-spike: detected (10.4 million)**<br><br>**Anti-nucleocapsid: detected (7.52)** | 190 days | 1st episode: symptomatic (mild, covid-19-like symptoms)<br><br>2nd episode: asymptomatic | 1st episode: Ct 36.0 (PHE RdRp assay)<br><br>2nd episode: Cn 21.2 (Abbott RdRp/N assay), repeat PCR on day 2 and day 4 both negative |
| HCW 3 | 52F, White | **Anti-spike: detected (8.8 million)**<br><br>Anti-nucleocapsid: not detected (0.02) | 199 days | 1st episode: symptomatic (mild)<br><br>2nd episode: asymptomatic when tested (transient myalgia following influenza vaccine previous week) | 1st episode: PCR-negative<br><br>2nd episode: Cn 12.6 (Abbott RdRp/N assay), repeat day 2 Ct 24.0 (Altona E/S gene) |
| HCW 4 | 26F, Asian | **Anti- spike: detected (11.6 million)**<br><br>**Anti-nucleocapsid: detected (3.9)** | 231 days | 1st episode: symptomatic (mild, fever and anosmia)<br><br>2nd episode: asymptomatic | 1st episode: not done (prior to start of symptomatic staff testing)<br><br>2nd episode: Ct 10 (Thermofisher, N gene assay), not repeated |

**Supplementary Table S5. Demographic, clinical and laboratory characteristics of individuals with possible SARS-CoV-2 reinfection.** *Time between episodes is calculated from date of positive PCR if index infection was PCR-positive (HCW 2 and 3), date of symptom onset if index infection was symptomatic but no was PCR performed (HCW 4), or date of first clinic attendance if presumed first episode was asymptomatic with no PCR performed (HCW 1).
